# Longitudinal plasma proteomics separates diagnostic differences from progression-linked changes in Alzheimer’s disease

**DOI:** 10.64898/2026.07.01.26356385

**Authors:** Junyoung Park, Yann Le Guen, Andrés Peña-Tauber, Michael D. Greicius

## Abstract

Most plasma proteomic studies in Alzheimer’s disease (AD) compare cases and controls cross-sectionally, leaving unresolved which AD-associated proteins mark diagnostic states and which are linked to disease progression. Using longitudinal SomaScan profiling from the Global Neurodegeneration Proteomics Consortium (13,449 participants, 17,269 samples, 7,362 aptamers), we separated baseline AD differences from AD-specific change over time. Linear mixed-effects models requiring concordant baseline and AD-by-time effects defined a 30-protein signature. We prioritized proteins across five evidence domains: clinical progression, AD biomarker alignment, cerebrospinal fluid concordance, independent prospective replication in UK Biobank and genetic support from Mendelian randomization and rare-variant burden. Thirteen proteins were supported in two or more domains and six in three. EDA2R, HPGDS, ITGAV and CLEC3B converged across clinical, biomarker and prospective evidence. Signature proteins aligned more strongly with tau and neuronal-injury markers than with A*β*42/40. ANTXR1 showed direction-concordant plasma pQTL Mendelian randomization and nominal rare-variant burden signals, supporting its prioritization within the longitudinal AD signature. By distinguishing diagnostic-state markers from progression-linked changes, this longitudinal, multi-domain approach prioritizes proteins for validation as markers of AD progression and for mechanistic and therapeutic follow-up.

---

Alzheimer’s disease (AD), the most common cause of dementia, develops over decades and contributes substantially to a global dementia burden, projected to rise from an estimated 57 million people in 2019 to more than 150 million by 2050.^1–3^ Accessible blood-based biomarkers are central to early detection, disease staging and trial enrichment, particularly with the recent approval of anti-amyloid therapies that require timely identification of patients in early symptomatic stages.^4–6^ Plasma proteomics extends beyond single-analyte readouts and enables simultaneous interrogation of thousands of proteins, capturing broader aspects of AD-associated biology including inflammation, vascular function and neural injury responses.^7,8^ Recent large-scale plasma proteomic studies using SomaScan and Olink platforms have identified AD-associated proteins in both case–control and prospective designs, supporting the plasma proteome as a resource for biomarker discovery.^9–11^ Beyond their biomarker utility, plasma protein signals may also identify disease-associated pathways and nominate candidates for mechanistic or therapeutic follow-up. These advances motivate systematic interpretation of plasma protein findings in relation to disease stage, progression and biological context.

Nevertheless, most large plasma proteomic studies in AD have focused on case–control associations at a single time point, identifying proteins that differ between diagnosed individuals and controls on cross-sectional panels such as SomaScan and Olink.^9–11^ In turn, longitudinal designs have typically been modest in cohort size^12^ or focused on a handful of established analytes, including plasma phosphorylated tau (pTau)-217, pTau-181 and amyloid-*β* 42/40 (A*β*42/40).^13–16^ A protein that is altered at baseline but stable over follow-up may mark a diagnostic state, whereas a protein that is altered at baseline and continues to diverge with AD status over time may better reflect a disease-associated trajectory. Thus, modeling both baseline and longitudinal changes in AD at proteome scale may nominate new proteins associated with disease progression.

Even when longitudinal modeling is performed, plasma proteomic discovery in AD can nominate dozens to hundreds of candidates whose biological origins are heterogeneous.^9,10^ Circulating protein levels may reflect central nervous system pathology, but also peripheral immune activity, vascular and metabolic state, assay-specific epitope behavior, or comorbid disease burden.^17^ For clinical and biological interpretation, longitudinal association alone may therefore be insufficient to distinguish progression-linked candidates from reactive or systemic correlates of disease status. A complementary approach is to prioritize candidates based on their signal across evidence domains spanning clinical progression, established disease biomarkers, external prospective cohorts, cerebrospinal fluid (CSF) concordance and genetic instruments. Under this multi-domain prioritization, signals supported by several domains are advanced as higher-confidence candidates, while single-domain hits are retained as discovery-level annotations pending further evidence.

We developed a longitudinal, multi-domain prioritization strategy for AD plasma proteomics and applied it to the Global Neurodegeneration Proteomics Consortium (GNPC) plasma SomaScan v4.1 resource, comprising 17,269 longitudinal samples from 13,449 participants across 7,362 aptamers. By modeling both baseline AD differences and AD-specific longitudinal change, then prioritizing candidates across five complementary evidence domains, we separated broad AD-associated plasma remodeling from a smaller set of convergently supported progression-linked candidates.

## Results

### Study design and multi-domain prioritization

To prioritize proteins with evidence beyond longitudinal association alone, we mapped the 30-protein signature across five complementary evidence domains, each addressing a distinct question (Figure 1). Clinical progression was tested in GNPC with Cox models of Clinical Dementia Rating (CDR)^18,19^ change over time. AD biomarker alignment was assessed against pTau-217, pTau-181, A*β*42/40, glial fibrillary acidic protein (GFAP) and neurofilament light chain (NfL) in the Stanford Alzheimer’s Disease Research Center (ADRC). External prospective replication used UK Biobank (UKB) Olink for incident AD and all-cause dementia. Cross-tissue concordance was examined in GNPC CSF. Genetic support was assessed using two independent lines of evidence: plasma pQTL Mendelian randomization (MR) with deCODE^20^ and UKB Pharma Proteomics Project (UKB-PPP)^21^ instruments, and rare-variant loss-of-function burden testing in a population-scale Alzheimer’s disease and related dementias (ADRD) proband meta-analysis of UK Biobank,^22^ All of Us,^23^ the Alzheimer’s Disease Sequencing Project (ADSP)^24^ and previously published Holstege case–control summary statistics.^25^ Convergence across these complementary domains served as the prioritization criterion, distinguishing broad AD-associated plasma remodeling from a focused set of progression-linked candidates.

**Figure 1.**
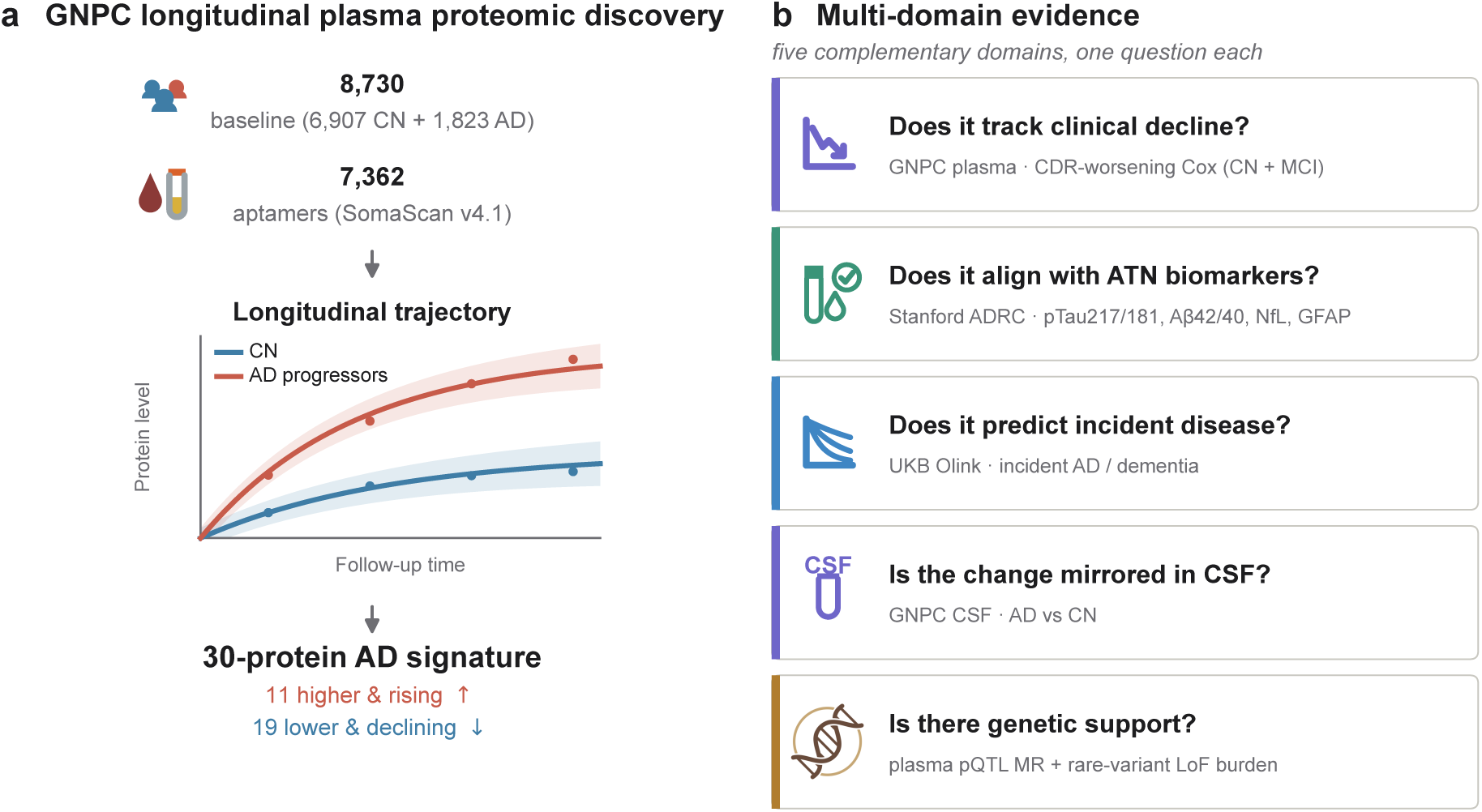
Study design for longitudinal plasma proteomic discovery and multi-domain prioritization in Alzheimer’s disease. **a**, GNPC plasma SomaScan v4.1 profiling included 13,449 participants, 17,269 longitudinal samples and 7,362 aptamers across 17 sites. Discovery was restricted to cognitively normal controls and AD participants (8,730 participants; 10,938 samples) and used mixed-effects models to identify proteins with both a baseline AD difference and AD-specific longitudinal change. The resulting 30-protein longitudinal AD signature comprised 11 Q1 proteins, defined by positive baseline AD and AD-by-time effects, and 19 Q3 proteins, defined by negative effects for both terms. **b**, Each signature protein was then evaluated across five complementary evidence domains: clinical progression using GNPC CDR-worsening Cox models; AD-related plasma biomarker alignment in Stanford ADRC across A*β*42/40, pTau-181, pTau-217, GFAP and NfL; external prospective replication in UKB Olink for incident AD and all-cause dementia; CSF concordance in GNPC CSF; and genetic support combining plasma pQTL Mendelian randomization (deCODE and UKB-PPP instruments) with rare-variant loss-of-function burden (ADRD proband meta-analysis of UK Biobank, All of Us, ADSP and published Holstege summary statistics^25^). **Abbreviations:** UKB-PPP, UK Biobank Pharma Proteomics Project; ADRD, Alzheimer’s disease and related dementias; ADSP, Alzheimer’s Disease Sequencing Project.

### A 30-protein plasma signature captures AD-specific longitudinal change

For each of the 7,362 SomaScan v4.1 aptamers, we fitted a linear mixed-effects model in the discovery subset (8,730 cognitively normal (CN) and AD participants, 10,938 samples) with two terms of interest. The baseline AD main effect estimated whether protein abundance differed between AD cases and cognitively normal controls at study entry. The AD-by-time interaction asked whether protein levels changed at a different rate in AD than in controls. Together these terms separate broad cross-sectional AD differences from the smaller set of proteins whose plasma levels track an AD-specific trajectory. We applied Benjamini–Hochberg false discovery rate (FDR) correction^26^ to each term independently across all aptamers.

The baseline AD main effect was widespread. At FDR < 0.05, 3,265 of the 7,362 aptamers differed between AD and control at baseline, 1,726 higher and 1,539 lower in AD (Figure 2a). Such breadth points to broad cross-sectional remodeling of the plasma proteome in AD. The AD-by-time interaction was far more selective. Only 86 aptamers showed a significant AD-specific change over time, 45 rising and 41 falling more steeply in AD (Figure 2b), which suggests that most baseline differences hold steady across follow-up rather than continuing to diverge.

**Figure 2.**
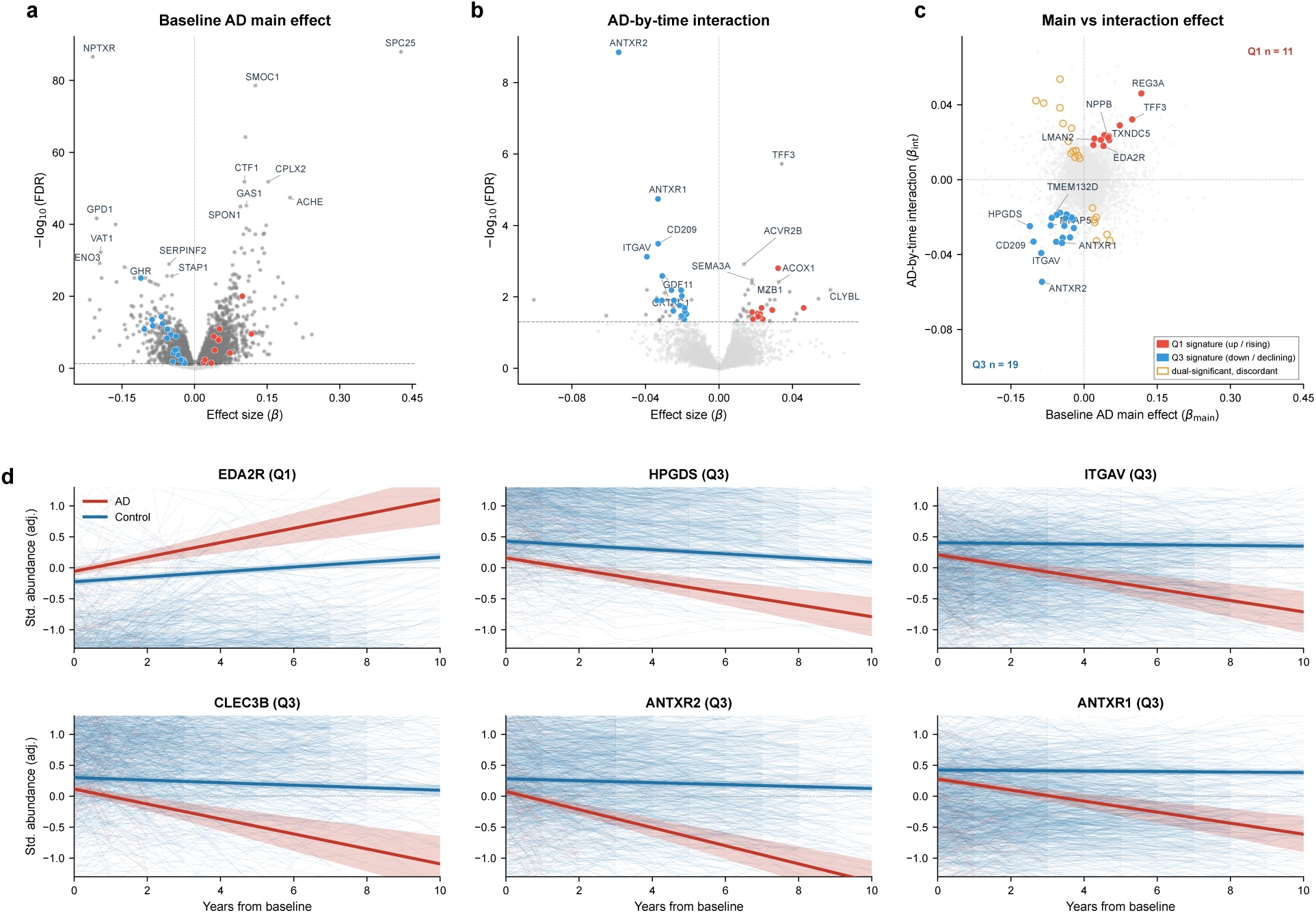
Discovery of a 30-protein longitudinal AD plasma signature. For each of 7,362 SomaScan v4.1 aptamers in the GNPC cognitively normal control and AD discovery subset, mixed-effects models tested two terms: the baseline AD main effect and the AD-by-time interaction. **a,** Volcano plot of the baseline AD main effect (AD-versus-control difference at baseline). **b,** Volcano plot of the AD-by-time interaction (whether protein levels changed differently over time in AD versus controls). In **a** and **b**, the *x* axis shows the estimated effect size and the *y* axis −log_#(_(FDR), with dashed lines at FDR = 0.05 and FDR controlled separately per term across all aptamers. Gray points did not pass term-specific FDR < 0.05, dark gray points passed but were not retained in the final signature, and red and blue points are the final Q1 and Q3 signature proteins. **c,** Quadrant plot integrating the two terms. Proteins passing FDR < 0.05 for both with concordant directions defined the signature: 11 Q1 proteins, higher in AD at baseline and rising over time, and 19 Q3 proteins, lower at baseline and declining. Light orange points are dual-significant proteins with discordant directions, not retained. **d,** Covariate-adjusted longitudinal trajectories for the six proteins supported in three evidence domains (EDA2R, HPGDS, ITGAV, CLEC3B, ANTXR2 and ANTXR1), standardized and adjusted for sex, baseline age, study site and the first five proteomic principal components. Bold lines show model-predicted AD (red) and control (blue) mean trajectories with shaded 95% confidence bands; faint lines show individual covariate-adjusted trajectories for participants with two or more plasma visits. Q1 proteins rise and Q3 proteins decline in AD over follow-up, while control trajectories stay near-flat; trajectories are shown to 10 years, beyond which fewer than 6% of samples contributed. Labeled points mark top-ranked aptamers by term-specific FDR in **a** and **b** and representative signature proteins in **c**.

Intersecting the two terms isolated proteins that were both altered at baseline and changed AD-specifically over time. Of the 52 aptamers significant for both terms, 21 had discordant effect directions, with the baseline AD difference and the AD-specific longitudinal change pointing opposite ways. We excluded these discordant aptamers because their baseline and longitudinal effects differed in direction. The retained sign-concordant set defined a 30-protein longitudinal AD plasma signature, roughly 0.4% of the SomaScan v4.1 platform. The signature split into two opposing patterns. The signature comprised 11 Q1 proteins, defined by positive baseline AD and AD-by-time effects, and 19 Q3 proteins, defined by negative effects for both terms (Figure 2c). Q1 proteins were higher in AD at baseline and diverged further in the positive direction over time, whereas Q3 proteins were lower in AD at baseline and diverged further in the negative direction.

Full estimates are in Supplementary Table 1. Covariate-adjusted model-predicted trajectories reproduced this pattern: Q1 proteins rose and Q3 proteins declined in AD relative to controls across follow-up, while control trajectories stayed near-flat (Figure 2d for the six proteins later supported in three evidence domains; Supplementary Figure 1 for the remaining signature proteins).

Sensitivity analyses supported the stability of the 30-protein signature after refitting the full-panel discovery model under alternative specifications (Supplementary Table 2). Using either a measured-CDR baseline diagnosis or restricting to sites contributing both control and AD participants left all 30 proteins concordant in direction, with effect-size estimates closely correlated with the primary model (Pearson *r* = 0.99–1.00 for the baseline effect and 0.90–1.00 for the interaction); the number of aptamers independently re-clearing both FDR thresholds varied with analytic scope and power rather than signal. Removing site adjustment expanded the signature to 87 proteins (29 of the 30 retained), reflecting uncorrected site and batch structure and supporting the site-adjusted model as primary. Adjusting for *APOE ε*2 and *ε*4 allele dose in the genotyped subset left all 30 estimates essentially unchanged (Pearson *r* = 0.997 baseline, 0.987 interaction), indicating the signature is not an *APOE* artifact.

### Convergent cross-domain evidence prioritizes 13 proteins

To prioritize candidates, we tested each of the 30 signature proteins across five evidence domains: clinical progression in GNPC plasma using Cox proportional hazards models for CDR worsening, AD-related biomarker alignment in the Stanford ADRC, CSF concordance in GNPC, external prospective replication in UKB Olink for incident AD and all-cause dementia, and genetic support from two complementary analyses: plasma pQTL Mendelian randomization and rare-variant loss-of-function burden in a population-scale ADRD meta-analysis. We tested each domain under its own pre-specified FDR family and counted a cross-domain association as supportive only when it reached FDR < 0.05 with a direction matching the AD longitudinal signature. For the genetic domain, a gene qualified when either plasma pQTL MR or rare-variant burden met this FDR-significant, direction-concordant bar; weaker nominal burden signals are reported separately as exploratory.

Figure 3a reports the number of directionally consistent proteins supported in each domain, separating FDR-significant associations from additional nominal ones. At FDR < 0.05, 9 proteins tracked subsequent CDR worsening in GNPC, 20 aligned with at least one of five AD-related biomarkers in Stanford ADRC, 5 showed a concordant AD direction in GNPC CSF, and 10 of the 18 panel-overlapping proteins replicated for incident AD or dementia in UKB. Nominal-only associations added exploratory support in several domains but did not enter the primary evidence count. The Stanford ADRC biomarker signal was tau-and injury-dominant. At FDR < 0.05, the per-biomarker counts were 18 for NfL, 8 for pTau-181, 8 for GFAP, 5 for pTau-217 and only 1 for A*β*42/40. One protein, ANTXR1, reached FDR < 0.05 in plasma pQTL MR, the only FDR-significant MR association; in rare-variant burden no gene survived FDR correction within the signature, though *ANTXR1*, *PAM* and *ATF6* carried direction-concordant burden at nominal significance.

**Figure 3.**
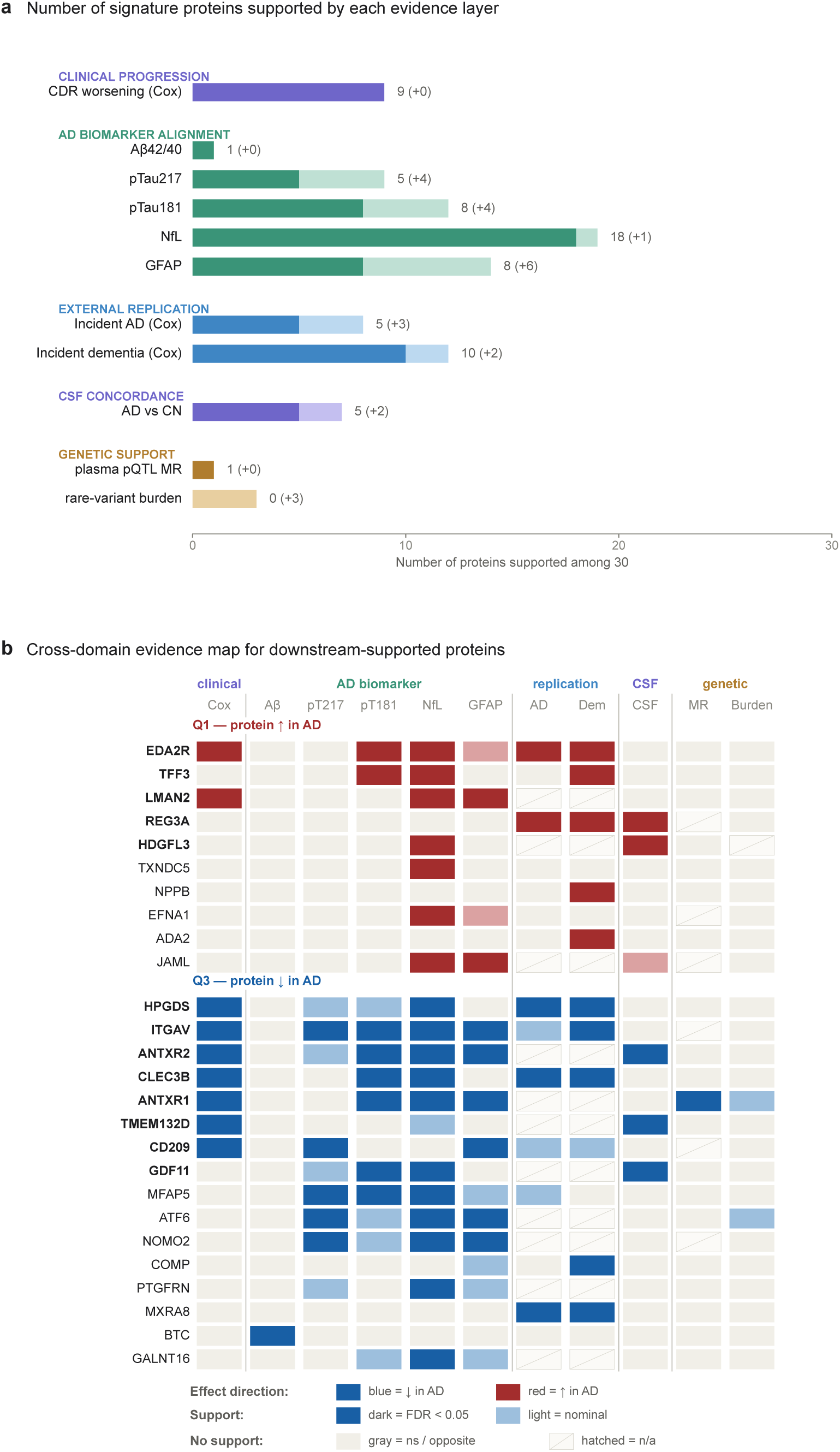
Multi-domain prioritization of the 30-protein longitudinal AD plasma signature. a, Counts of signature proteins with FDR-significant and direction-consistent support across five evidence domains: clinical progression in GNPC plasma, AD-related plasma biomarker alignment in Stanford ADRC, external prospective replication in UK Biobank Olink, CSF concordance in GNPC, and genetic support, the last shown as two bars, plasma pQTL Mendelian randomization and rare-variant loss-of-function burden in an ADRD proband meta-analysis. Dark bars indicate associations passing FDR < 0.05 within the corresponding domain-specific testing family and matching the Q1/Q3 discovery direction; light extensions indicate additional direction-consistent nominal associations at *p* < 0.05 that did not pass FDR. For rare-variant burden, FDR was computed within the 30-gene family per ancestry, under a loss-of-function model in which reduced gene product raises risk for AD-declining proteins. **b,** Cross-domain evidence map for the 26 signature proteins with at least one FDR-significant, direction-consistent cross-domain association (10 Q1, 16 Q3). Rows represent proteins and columns represent evidence domains. Tile color indicates the discovery-aligned direction of support, with red for Q1-aligned effects and blue for Q3-aligned effects. Tiles are colored only where the domain effect is direction-consistent with the protein’s discovery direction; for the A*β*42/40 ratio, which decreases in AD, consistency is defined relative to that decrease. Dark tiles indicate FDR-significant support, light tiles indicate nominal direction-consistent support, gray tiles indicate tested associations that were non-significant or directionally discordant, and hatched tiles indicate proteins or tests unavailable in that domain. The two genetic columns show plasma pQTL MR and rare-variant burden; because loss-of-function and protein-abundance effects can diverge, discordant genetic signals appear gray and are reported in the text. Thirteen proteins were supported in two or more evidence domains and were defined as the multi-domain subset; six of these were supported in three domains. Full cross-domain effect sizes, *p* values and FDR values for all 30 signature proteins are provided in Supplementary Table 3.

Figure 3b summarizes the evidence domains supporting each protein. Of the 30 longitudinal signature proteins, 26 had at least one FDR-significant, direction-consistent cross-domain association, including 10 Q1 and 16 Q3 proteins. Convergent support was seen in a smaller subset. Thirteen proteins were supported in two or more evidence domains, comprising five Q1 proteins (EDA2R, TFF3, REG3A, HDGFL3 and LMAN2) and eight Q3 proteins (HPGDS, ITGAV, ANTXR2, ANTXR1, CLEC3B, CD209, TMEM132D and GDF11). We refer to these 13 proteins, hereafter, as the multi-domain subset.

Within this subset, six proteins drew support in three domains. Four of them (EDA2R, HPGDS, ITGAV and CLEC3B) were supported across clinical progression, Stanford ADRC biomarker alignment and UKB prospective replication. ANTXR2 was supported by clinical progression, Stanford ADRC biomarker alignment and CSF concordance but was not assayed on the UKB Olink panel. ANTXR1 was supported by clinical progression, Stanford ADRC biomarker alignment and genetic evidence, with signals in both plasma pQTL MR and direction-concordant rare-variant burden. The remaining seven convergently supported proteins were supported in two domains, with evidence combinations shown in Figure 3b and Supplementary Table 3. The multi-domain subset is therefore best read as a structured follow-up set with heterogeneous evidence patterns rather than a set of equivalent candidates. Supplementary Table 3 gives the full cross-domain effect-size and FDR matrix for all 30 proteins.

### Signature proteins are associated with subsequent CDR worsening in GNPC

We first asked whether the 30-protein signature in GNPC plasma tracked subsequent clinical progression. We fitted Cox proportional hazards models for all 30 signature aptamers in CN and MCI participants (the primary scope; 1,762 participants, 156 CDR-worsening events), adjusting for baseline age, sex, baseline CDR category, study site and the first five plasma proteomic principal components, with Benjamini–Hochberg correction across the 30 aptamers. Nine proteins reached FDR < 0.05 with direction-consistent effects.

The nine associations spanned both discovery quadrants, reported as hazard ratios (HR) per 1 s.d. higher baseline abundance. Two Q1 proteins (EDA2R, LMAN2) were associated with higher hazard of subsequent CDR worsening, and seven Q3 proteins (HPGDS, CLEC3B, CD209, TMEM132D, ITGAV, ANTXR2 and ANTXR1) with lower hazard; the strongest were EDA2R (HR = 1.47) and HPGDS (HR = 0.68). Higher Q1 abundance and lower Q3 abundance marked greater risk of worsening, matching the discovery trajectories (Figure 4a; full hazard ratios, confidence intervals and FDR values in Supplementary Table 4). All nine proteins were members of the multi-domain subset.

**Figure 4.**
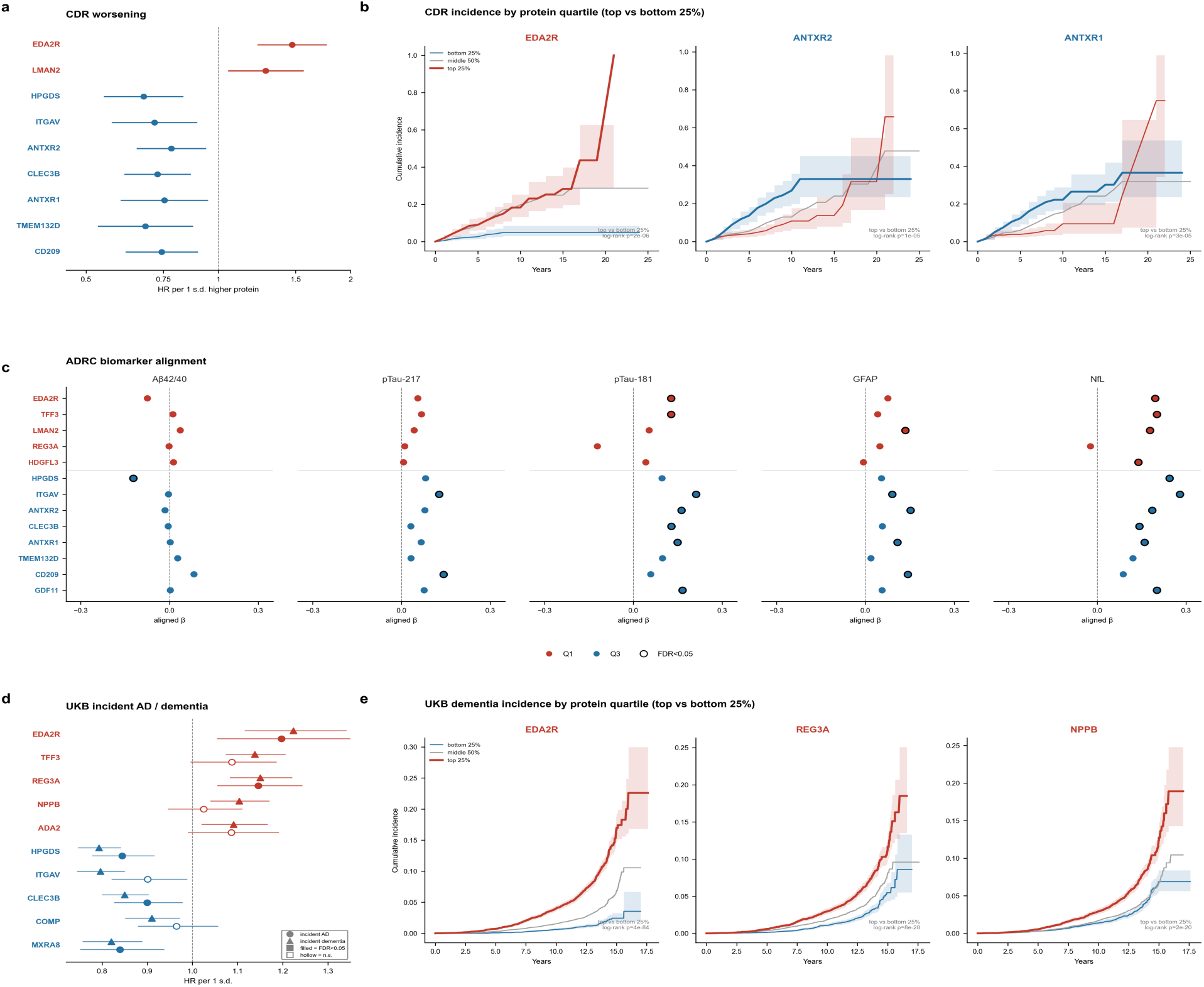
Effect patterns across clinical progression, AD biomarker alignment and prospective replication. a, Cox proportional hazards estimates for the nine signature proteins with FDR-significant, direction-consistent associations with CDR worsening in GNPC plasma, as hazard ratios (HR) per 1 s.d. higher baseline abundance with 95% confidence intervals; the vertical line marks HR = 1. **b,** Representative cumulative incidence curves for CDR worsening in GNPC, with participants split into bottom 25%, middle 50% and top 25% of raw baseline abundance (visualization only; inference used standardized continuous abundance) and a log-rank contrast between the top and bottom quartiles; the AD-like direction is the top quartile for Q1 proteins and the bottom quartile for Q3 proteins. **c,** Stanford ADRC biomarker associations for the 13 multi-domain subset proteins across A*β*42/40, pTau-217, pTau-181, GFAP and NfL; dot color is the standardized effect aligned to the AD-like discovery direction and a dark outline marks FDR < 0.05 under the combined 145-test family. **d,** UKB Olink Cox estimates (HR per 1 s.d.) for the ten signature proteins with FDR-significant, direction-consistent incident-dementia replication, shown for incident AD and all-cause dementia; proteins absent from the Olink panel are not plotted. **e,** Representative cumulative incidence curves for incident dementia in UKB, stratified as in **b**. Representative proteins in **b** and **e** were chosen among FDR-significant, direction-consistent Cox associations, prioritizing multi-domain subset proteins and spanning both discovery directions. Red and blue denote Q1 and Q3 assignment, respectively.

Because *APOE* is the major common genetic risk locus for AD, we tested whether these associations were sensitive to *APOE* adjustment. In the genotyped subset (1,085 participants, 63 events), adding *APOE ε*2 and *ε*4 allele dose left the nine hazard ratios essentially unchanged (ratio of *APOE*-adjusted to unadjusted HR, 0.99–1.01; Supplementary Table 4); the smaller event count precluded its use as the primary model, so it is reported as a sensitivity analysis.

Quartile-stratified cumulative incidence curves (top versus bottom 25% of baseline abundance) separated CDR-worsening risk in the expected direction for representative Q1 and Q3 proteins, agreeing with the Cox estimates (Figure 4b).

### Signature proteins align with AD biomarker burden in Stanford ADRC

To test whether the longitudinal plasma signature aligned with established AD-related plasma biomarker axes, we evaluated each signature protein against biomarker measurements in the Stanford ADRC within GNPC. The Stanford ADRC analytic set held 293–302 unique participants with longitudinal plasma biomarker assays, with a median of 534 observations per protein–biomarker model. We estimated protein–biomarker associations using linear mixed-effects models with a participant-level random intercept to account for repeated measurements. Twenty-nine of the 30 signature aptamers were present on the Stanford ADRC SomaScan panel, and we tested each against five AD-related plasma biomarkers spanning amyloid, tau and injury/glial pathways: A*β*42/40, pTau-181, pTau-217, GFAP and NfL. Benjamini–Hochberg correction across 145 tests controlled multiple testing.

At FDR < 0.05 with direction-consistent effects, the per-biomarker hit counts across the full signature were 18 for NfL, 8 for pTau-181, 8 for GFAP, 5 for pTau-217 and only 1 for A*β*42/40 (Supplementary Table 5). Statistical power is an unlikely explanation for these differences, because the number of contributing observations stayed comparable across the five biomarkers (534–585). Alignment thus clustered on tau-related and injury/glial markers, while amyloid associations stayed sparse. A *β* 42/40 produced one direction-consistent FDR-significant association across the entire 29-aptamer signature, BTC, which was otherwise not part of the multi-domain subset.

The same tau-and injury-dominant pattern held within the multi-domain subset in the Stanford ADRC biomarker dot plot (Figure 4c). Eight of the 13 subset proteins reached FDR < 0.05 for pTau-181 or pTau-217. The largest single effect belonged to ITGAV with pTau-181 (*β* = −0.21, *p* = 6.9 × 10^-6^), which also held a consistent direction across all four tau and injury/glial markers (pTau-181, pTau-217, GFAP and NfL). None of the 13 subset proteins reached FDR < 0.05 for A*β* 42/40. Other subset proteins, among them REG3A and TMEM132D, returned no FDR-significant Stanford ADRC biomarker alignment and instead drew support from clinical progression, CSF concordance or external replication.

### UK Biobank Olink supports prospective associations with incident AD and dementia

To test the 30-protein signature for prospective replication, we used the UKB-PPP Olink Explore panel, which measured 2,923 plasma proteins in 54,219 UKB participants.^21^ After merging data and removing participants with baseline-prevalent disease, 38,243 remained for incident AD (622 events) and incident all-cause dementia (1,171 events). Eighteen of the 30 signature aptamers had a matching Olink analyte (Supplementary Table 6). Cox models adjusted for baseline age, sex and the first five Olink proteomic principal components. Benjamini–Hochberg correction spanned 36 tests.

At FDR < 0.05 with direction-consistent effects, 5 proteins replicated for incident AD and 10 for incident all-cause dementia, the better-powered endpoint. The two strongest were both Q3 proteins for incident dementia, HPGDS (HR = 0.79, 95% CI 0.75–0.84; *p* = 1.1 × 10^-14^) and ITGAV (HR = 0.80; *p* = 5.0 × 10^-12^) (Figure 4d).

Among the 13 multi-domain subset proteins, 7 were on the UKB Olink panel and 6 replicated for at least one outcome (CLEC3B, EDA2R, HPGDS, ITGAV, REG3A and TFF3); the other six (ANTXR1, ANTXR2, GDF11, HDGFL3, LMAN2 and TMEM132D) were not assayed. Across the two outcomes, replicated Q1 proteins (EDA2R, REG3A, TFF3, NPPB and ADA2) showed higher risk and Q3 proteins (HPGDS, ITGAV, CLEC3B, MXRA8 and COMP) lower risk; EDA2R, REG3A, HPGDS, CLEC3B and MXRA8 replicated for both incident AD and dementia. Per-protein hazard ratios, confidence intervals and FDR values are in Supplementary Table 6.

Quartile-stratified cumulative incidence curves for incident dementia separated in the expected direction (high-abundance quartile for Q1 proteins, low-abundance for Q3), agreeing with the Cox estimates (Figure 4e).

### CSF and genetic analyses support selected progression-linked proteins

To ask whether the plasma-derived signature carried across tissues, we tested GNPC CSF proteomics in CN and AD participants, regressing CSF protein levels on AD status with the same covariates. Of 7,351 CSF aptamers, 1,103 differed by AD status at FDR < 0.05. Among the 30 plasma signature aptamers, 5 reached FDR < 0.05 in CSF, all in the same direction as the plasma signature and all members of the multi-domain subset: GDF11, TMEM132D, HDGFL3, REG3A and ANTXR2 (Supplementary Table 7). For GDF11 and HDGFL3, CSF concordance paired with Stanford ADRC biomarker alignment, and for ANTXR2 it was the third supporting domain. CSF support was otherwise sparse: the other nine subset proteins lacked direction-consistent CSF signal, consistent with cross-tissue heterogeneity between plasma and CSF.

For a genetic-support domain, we ran two-sample MR using plasma pQTL instruments from deCODE and UKB-PPP under strict aptamer-identifier matching. One protein, ANTXR1 (Q3; inverse-variance weighted *β* = −0.39, FDR = 0.046 from two cis instruments), reached FDR < 0.05 (Supplementary Table 8). Colocalization across the *ANTXR1* cis locus, however, gave only a 0.6% posterior probability of a shared causal variant (PP.H4), against 53% for a pQTL-only signal, and regional AD association stayed weak; the cis-MR estimate is therefore not corroborated by colocalization.

As a second genetic line of evidence, we evaluated all 30 longitudinal signature genes in a population-scale ADRD rare-variant burden meta-analysis of high-confidence loss-of-function variants, using European-ancestry (Supplementary Table 9). No gene survived FDR correction within the 30-gene family, but ANTXR1 showed the strongest direction-concordant burden signal (Q3; OR = 6.75, P = 0.0025, FDR = 0.063), with PAM (OR = 2.10, P = 0.0045, FDR = 0.063) and ATF6 (OR = 2.08, P = 0.032, FDR = 0.227) also reaching nominal significance in the expected direction. ANTXR1 was therefore the only signature gene with both an FDR-significant plasma pQTL MR association and a direction-concordant nominal rare-variant burden signal. REG3A showed nominal rare-variant burden in the direction opposite to its Q1 plasma trajectory, consistent with divergence between loss-of-function effects and circulating protein-abundance associations.

## Discussion

Most plasma proteomic studies of AD compare cases and controls at a single time point, identifying which proteins differ but not how their abundance changes as disease progresses.^9,10^ We addressed this gap by requiring both a baseline AD difference and an AD-specific longitudinal change, thereby prioritizing proteins linked to disease-associated plasma remodeling over time rather than fixed cross-sectional differences. In the CN and AD discovery subset of GNPC, this dual-term model yielded a 30-protein longitudinal AD plasma signature, comprising 11 Q1 proteins that were higher in AD and increased over time and 19 Q3 proteins that were lower in AD and decreased over time.

The cross-domain evidence was heterogeneous rather than uniformly concordant, which we regard as an expected feature of plasma biomarker discovery rather than a weakness: a plasma scan captures broad disease-associated remodeling that includes systemic, inflammatory, vascular and reactive components alongside signals tied to clinical progression. Across five complementary domains, 26 of the 30 proteins had at least one FDR-significant, direction-consistent cross-domain association, 13 in two or more domains (the multi-domain subset) and six in three. Any single association may reflect reactive, systemic or cohort-specific biology, whereas agreement across complementary outcomes, tissues, assays, an external cohort and genetic instruments is unlikely to arise from one artefact; convergence therefore prioritizes proteins more confidently linked to disease.

The 13 multi-domain subset proteins should not be interpreted as a single ranked list. The evidence domains differed in the type of support they provided: Stanford ADRC biomarker alignment and CSF concordance reflected molecular associations, CDR worsening and UKB incident-disease analyses reflected clinical or prospective outcome associations, and plasma pQTL MR and rare-variant burden provided genetic support. The pattern of evidence for each protein is therefore more informative than the number of supporting domains alone. Proteins supported by clinical, biomarker and prospective evidence should be interpreted differently from ANTXR1, whose convergent support included genetic evidence.

The strongest convergent pattern came from four proteins (EDA2R, HPGDS, ITGAV and CLEC3B) supported across clinical progression, AD biomarker alignment and external prospective replication, the most consistently replicated tier of the study. Together with the other subset proteins, these candidates are consistent with neuroinflammation, extracellular-matrix and vascular remodeling, and systemic aging-related stress as biological contexts for mechanistic validation and therapeutic follow-up.

Several proteins point to inflammatory and glial biology. The clearest is HPGDS (Q3), which encodes hematopoietic prostaglandin D synthase; HPGDS and the DP1 receptor have been localized to microglia and astrocytes within senile plaques in human AD and an AD mouse model.^27^ Lower HPGDS abundance was part of the AD-like longitudinal pattern and was associated with faster CDR worsening in GNPC and higher incident-dementia risk in UKB, the strongest such association in the signature, so HPGDS may index a microglial or prostaglandin-related inflammatory state, although plasma data cannot distinguish disease response from compensatory or protective biology.

A larger group implicates extracellular-matrix, adhesion and vascular biology. ITGAV (Q3) encodes integrin *α*V, a cell–matrix adhesion and TGF-*β*-activating receptor subunit implicated in amyloid-*β*-induced neurotoxicity;^28^ it replicated for incident dementia and showed the strongest pTau-181 alignment of any signature protein. CLEC3B (Q3) encodes tetranectin, a plasminogen-binding lectin involved in proteolysis and tissue remodeling whose abundance is reduced in CSF across neurological disease.^29,30^ The two anthrax toxin receptors ANTXR1 (TEM8) and ANTXR2 (CMG2), both Q3, bind collagen and participate in matrix homeostasis and angiogenesis;^31–34^ ANTXR1 was the only signature protein with both a plasma pQTL MR association and direction-concordant rare-variant burden (loss-of-function OR = 6.7, P = 0.0025), although its MR estimate did not colocalize with AD risk and the burden was nominal, so its genetic support is suggestive rather than conclusive. Together these proteins are consistent with, but do not establish, an extracellular-matrix and vascular component of the longitudinal signature.

Two further proteins more likely reflect systemic than AD-specific biology and should be read cautiously. EDA2R (Q1, the strongest Q1 representative) is a ubiquitous hallmark of aging that mediates parainflammatory (chronic, low-grade inflammatory) responses,^35^ so its rise in AD may capture an aging-related component of the plasma trajectory rather than a CNS-specific mechanism. NPPB (Q1) encodes B-type natriuretic peptide, a cardiac-stress hormone; elevated natriuretic peptides associate with vascular brain injury and dementia risk,^36^ matching its direction here, although a SomaScan study reported lower BNP in AD,^37^ so NPPB more plausibly indexes a cardiovascular or comorbidity readout than an AD-specific process.

Alignment with established AD biomarkers in Stanford ADRC was tau-and injury-dominant: across 29 aptamers tested against five biomarkers (145 tests), 18 proteins were FDR-significant and direction-consistent for NfL, 8 for pTau-181, 8 for GFAP, 5 for pTau-217 and only one for A*β*42/40. This stronger alignment with tau and injury or glial markers than with A*β*42/40 suggests the signature tracks downstream neurodegenerative and inflammatory processes more than amyloid burden itself; because plasma pTau rises largely in response to amyloid pathology, this does not exclude an amyloid-proximal process.^15,38^

The clinical and external prospective domains carry different caveats. Within GNPC, nine proteins reached FDR < 0.05 for CDR worsening; the UKB Olink analysis then provided external prospective support in a different population, platform and outcome definition, with six of seven UKB-testable subset proteins replicating for at least one outcome. This strengthens generalizability, but only 18 of the 30 SomaScan proteins were assayable on Olink, so absence from UKB often reflects lack of panel coverage rather than negative evidence, and incident all-cause dementia is etiologically heterogeneous.

CSF and MR illustrate the limits of cross-tissue and causal triangulation. CSF concordance was partial (5 of 30 proteins, all direction-consistent and all in the subset) but decisive for some: GDF11 and HDGFL3 paired CSF with ADRC alignment and ANTXR2 relied on CSF as its third domain. Because ADRC alignment and CSF concordance are both molecular domains, proteins supported only by this pair are not equivalent to those with clinical or prospective support. The incomplete plasma–CSF overlap indicates that plasma signatures are systemic and compartment-dependent rather than direct CSF surrogates.^39^

The genetic domain was deliberately conservative. Plasma pQTL MR yielded a single FDR-significant, direction-consistent association (ANTXR1) that did not colocalize with AD risk, and rare-variant burden added a second independent line in which ANTXR1, PAM and ATF6 carried nominal direction-concordant burden while REG3A diverged. Under strict aptamer-matched instrument selection, cis-pQTL coverage is incomplete and single-instrument MR cannot provide reliable triangulation, so the genetic domain functions as a complementary annotation rather than a causal selection gate.^40,41^

Several limitations should guide the interpretation of these results. First, the design is observational, so an AD-associated protein change may reflect causal contribution, downstream consequence, systemic response or residual confounding. Second, the clinical-progression, Stanford ADRC biomarker and CSF domains share participants with the discovery set and so constitute internal cross-domain support rather than sample-independent replication, with the UKB Olink and genetic analyses providing the externally independent axes. Third, platform differences between SomaScan v4.1 and UKB Olink limit cross-platform comparison, only 18 of 30 proteins overlap, and CSF sampling was cross-sectional, so longitudinal plasma–CSF coupling could not be assessed. Finally, the biological interpretations are literature-derived and were not tested mechanistically here, and the 13-protein multi-domain subset is a prioritization output, not an optimized clinical prediction panel, requiring prospective validation in cohorts with harmonized plasma, CSF, imaging and cognitive outcomes before any clinical utility can be assessed.^2,5^

We define a 30-protein longitudinal AD plasma signature and show that multi-domain evidence integration can prioritize proteins with convergent clinical-progression, AD-related biomarker, CSF and external prospective support. Rather than nominating a single definitive causal biomarker, our findings distinguish broad AD-associated plasma remodeling from progression-linked candidates and read those candidates by the pattern of evidence that supports them rather than by a single ranked score. Future studies should test these candidates in independent longitudinal cohorts and determine whether they reflect causal mechanisms, systemic responses or clinically useful markers of AD progression.

## Methods

### Data access and ethics

This study used the Global Neurodegeneration Proteomics Consortium (GNPC) Health Data Stewardship (HDS) v1 dataset, accessed through the Alzheimer’s Disease Data Initiative AD Workbench under the GNPC Data Use Agreement and publication policies.^11^

All contributing studies obtained local ethics committee or institutional review board approval and written informed consent from participants or their legal representatives at the site of origin.^11^ The present study was a secondary analysis of de-identified, harmonized data released through the GNPC HDS framework and did not constitute new human-subjects research at the analyzing institution.

### GNPC plasma cohort and sample selection

The primary plasma dataset was derived from the GNPC SomaScan v4.1 release, which assayed 7,362 aptamers across 17 contributing sites.^11^ We restricted this to EDTA plasma biological study samples, yielding a longitudinal plasma cohort of 17,269 samples from 13,449 participants (Table 1).

**Table 1.** Baseline characteristics of the GNPC plasma cohort. Continuous variables are mean (s.d.) or median [IQR]; categorical variables are *n* (%), summarized at each participant’s first plasma sample. *Note.* Diagnosis was based on global CDR, with a cognitive-battery fallback when CDR was missing; CN, MCI, and AD do not sum to the total because non-AD and unclassified diagnoses are omitted. *The signature’s associations were robust to *APOE* adjustment (ε2 and ε4 allele dose), as detailed in Results and Methods. *APOE ε*4 status was computed among 10,304 genotyped participants. ^a^Follow-up was summarized among participants with ≥2 plasma samples (*n* = 2,116). **Abbreviations:** AD, Alzheimer’s disease; CDR, Clinical Dementia Rating; CN, cognitively normal; IQR, interquartile range; MCI, mild cognitive impairment; MMSE, Mini-Mental State Examination; s.d., standard deviation.

| Characteristic | GNPC plasma cohort |
| --- | --- |
| Age, mean (s.d.) | 70.0 (12.9) |
| Female, *n* (%) | 7,431 (55.3%) |
| MMSE, mean (s.d.) | 23.8 (9.7) |
| APOE ε4 carrier*, *n* (%) | 3,666 (35.6%) |
| Cognitively normal, *n* (%) | 6,907 (51.4%) |
| Mild cognitive impairment, *n* (%) | 3,459 (25.7%) |
| Alzheimer's disease, *n* (%) | 1,823 (13.6%) |
| Visit 2, *n* (%) | 2,116 (15.7%) |
| Visit 3, *n* (%) | 1,249 (9.3%) |
| Visit $\geq 4$, *n* (%) | 421 (3.1%) |
| Follow-up duration (yr), median [IQR]<sup>a</sup> | 4.0 [2.0–7.0] |

For longitudinal discovery, the analytic set was limited to participants whose baseline diagnosis (the diagnosis recorded at each participant’s first visit) was cognitively normal (CN) or AD using the harmonized diagnosis variable described below, yielding 8,730 participants (6,907 controls and 1,823 AD cases) and 10,938 plasma samples. To reduce diagnostic inconsistency in the discovery analysis, CN with the global Clinical Dementia Rating (CDR)^18,19^ ≥ 1 and AD-labeled samples with CDR ≤ 0.5 were excluded before model fitting. Participants with baseline mild cognitive impairment (MCI) were excluded from the discovery linear mixed-effects model (LMM) but retained for the CDR-worsening clinical progression analysis, whereas those with non-AD neurodegenerative diagnoses or unclassified status were excluded from both the discovery LMM and the Cox analysis.

Repeated sampling in the discovery set was sparse: participants contributed a median of one plasma visit, with 14.7% providing two or more (maximum five). Diagnostic composition was uneven across contributing sites, and several sites contributed CN only (Supplementary Table 10). This structure motivated a participant-level random intercept without participant-specific random slopes, which are unidentifiable with one to two visits per participant, and the inclusion of contributing site as a model covariate.

### Diagnosis harmonization and clinical variables

GNPC pools cohorts spanning AD, Parkinson disease, frontotemporal dementia, amyotrophic lateral sclerosis and aging controls, each with distinct diagnostic workflows and cognitive assessment batteries. To place these data on a common clinical scale, we harmonized disease status into four categories: CN, MCI or subjective cognitive impairment (SCI), AD, and non-AD neurodegenerative disease.

The CDR was used whenever available. When CDR was missing, an equivalent CDR category was assigned from cognitive screening scores using prespecified mappings: Mini-Mental State Examination (MMSE)^42^ scores of 27–30, 21–26, 11–20 and 0–10 were mapped to CDR-equivalent categories of 0, 0.5, 1 and 2, respectively; and Montreal Cognitive Assessment (MoCA)^43^ scores of 26–30, 18–25, 11–17 and 0–10 were mapped to the same categories.

Diagnostic groups were assigned by harmonized clinical severity. CDR = 0 defined CN, CDR = 0.5 defined MCI or SCI, and CDR ≥ 1 defined AD unless cohort diagnosis indicated a non-AD neurodegenerative disorder. When CDR was unavailable, cohort-provided diagnosis labels were used, with control status assigned only when no neurodegenerative disease flag was present and the available clinical information was consistent with normal cognition. Cognitive status was treated as non-reverting across repeated visits: once a participant was classified as MCI, SCI or AD, a later normal CDR did not return the label to control, although progression to a more severe category was allowed.

### Plasma proteomic preprocessing and quality control

SomaScan aptamer relative fluorescence units^7^ were log2(*x* + 1) transformed to stabilize variance across the dynamic range. Quality-control steps followed the preprocessing pipeline of the primary GNPC proteomics study.^17^ Per-aptamer Tukey outliers^44^ falling outside [Q1 − 1.5 × IQR, Q3 + 1.5 × IQR] were set to missing. Call-rate filtering was then applied to both samples and aptamers in two successive rounds, first at 65% and then at 85% completeness.

Remaining missing values were imputed by k-nearest-neighbors with,*k* = √*n*^45^ after z-scoring each aptamer before the neighbor search and back-transforming imputed values to the log2 scale. Principal components were computed on the imputed log2 matrix, and the first five components (PC1–PC5) were included as technical covariates in the discovery mixed model and the CDR-worsening Cox model to adjust for residual platform and batch structure. CSF aptamer profiles were preprocessed using a parallel, independent pipeline.

Each aptamer was annotated with its UniProt accession^46^ and protein target from the SomaScan analyte table, and signature proteins are reported by UniProt accession alongside gene symbol; matching to UKB Olink assays used the shared UniProt target. Two signature aptamers map to more than one accession: the ITGAV aptamer targets the integrin *α*V*β*3 complex (UniProt P06756 and P05106), and the GDF11 aptamer cross-reacts with its homolog myostatin (MSTN; UniProt O95390 and O14793).

### Longitudinal plasma discovery by mixed-effects modeling

Longitudinal protein abundance was modeled by linear mixed-effects regression in R (version 4.3)^47^ using lme4^48^ with the lmerTest extension,^49^ which provides *p* values using Satterthwaite’s degrees-of-freedom approximation. Models were fitted to the discovery set defined above.

For each of the 7,362 aptamers, the per-aptamer preprocessed log2 abundance at each visit was the model outcome (not further standardized, so coefficients are reported on the log2 scale), modeled as a function of baseline diagnosis, time since the baseline plasma visit, and their interaction. Time (*t*, in years) was measured from each participant’s first plasma sample and coded so that *t* = 0 at the baseline visit; consequently the baseline AD main effect estimates the AD-versus-control difference at *t* = 0. Baseline diagnosis was defined as each participant’s diagnosis at their first (earliest sequential) plasma visit, and the discovery set was restricted to participants who were control or AD at that first visit; it was coded as AD versus control, with control as the reference group. Models adjusted for sex, baseline age, study site and the first five plasma proteomic principal components, and included a participant-level random intercept. The preprocessing pipeline and the proteomic principal components were derived from the full GNPC plasma matrix (all 17,269 samples across diagnostic groups) before restriction to the discovery set. Because protein abundance was the modeled outcome, the discovery model used the observed outlier-masked log2 values without imputation and fitted each aptamer on all available observations (available-case under a missing-at-random assumption); k-nearest-neighbor imputation was used only where a complete matrix was required, namely for the principal components and for the standardized baseline protein predictor in the downstream survival and biomarker analyses. The first five components were only weakly associated with baseline AD status (area under the curve 0.53–0.61 across PC1–PC5, taking the better-discriminating direction for each component), indicating that this full-cohort technical adjustment did not substantially absorb the AD signal. In compact form,

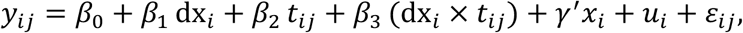

where dx_i_ is the baseline AD indicator, *t*_i,j_ is time since baseline, *x*_i_ denotes the covariates, and *u*_i_ is the participant-level random intercept. The two coefficients of interest were *β*_1_, representing the baseline AD-versus-control difference, and *β*_3_, representing the AD-specific difference in longitudinal change.

Multiple testing was controlled using the Benjamini–Hochberg procedure applied independently to each model term across all 7,362 aptamers, yielding term-specific false discovery rate (FDR)^26^ values for *β*_1_ and *β*_3_. The baseline and interaction terms were treated as distinct testing families, because they address different questions (a cross-sectional baseline AD difference and an AD-specific longitudinal divergence) with markedly different signal densities; per-term control prevents the abundant baseline signals from relaxing the threshold for the sparser interaction term. The longitudinal discovery signature required both terms to pass FDR < 0.05 and to show concordant effect directions. Aptamers with *β*_1_ > 0 and *β*_3_ > 0 were assigned to Q1, representing higher baseline abundance in AD with further increase over time. Aptamers with *β*_1_ < 0 and *β*_3_ < 0 were assigned to Q3, representing lower baseline abundance in AD with further decrease over time. This criterion excluded aptamers showing only a baseline difference or only an AD-specific temporal change without concordant direction.

### Sensitivity analyses

To assess the robustness of the 30-protein signature, we refit the discovery linear mixed-effects model under four alternative specifications and compared the baseline and AD-by-time effect estimates for the signature proteins against the primary model. First, the contributor (site) fixed effect was removed (no site adjustment). Second, baseline diagnosis was redefined from each participant’s measured baseline CDR (control, CDR 0; AD, CDR ≥ 1) rather than the harmonized diagnostic label. Third, the analysis was restricted to sites that contributed both control and AD participants. Fourth, as a confounding control, we adjusted for *APOE ε*2 and *ε*4 allele dose, each coded as the number of copies (0, 1 or 2), since *APOE* is the strongest common genetic risk locus for late-onset AD;^50,51^ both dose terms were added as fixed-effect covariates, not as interaction terms. *APOE* genotype was available for 10,304 of 13,449 participants (76.6%); participants without genotype were excluded and *APOE* status was not imputed.

Each model used per-term Benjamini–Hochberg FDR as in the discovery analysis. Stability was assessed from the Pearson correlation and the sign concordance of the signature proteins’ effect estimates between each model and the primary model, recognizing that the number of proteins that independently meet both FDR thresholds also depends on each model’s sample size. Full per-protein estimates under every model are reported in Supplementary Table 2.

### Clinical progression analysis using CDR worsening

CDR worsening was defined as the first increase in computed CDR after the baseline plasma visit. Follow-up time was measured from the baseline plasma draw to the first CDR increase, with participants censored at their last available CDR assessment if no worsening occurred. The primary eligible subset comprised participants with baseline CDR ≤ 0.5 (cognitively normal or MCI), yielding 1,762 participants and 156 incident worsening events; baseline CDR was modeled as CDR = 0 versus CDR = 0.5, with CDR = 0 as the reference. Two further scopes were prespecified as sensitivity analyses: a CN-only stratum (baseline CDR = 0) and a CN+MCI+AD scope in which CDR ≥ 1 was added as a third baseline severity category.

For each of the 30 signature aptamers, a Cox proportional hazards model^52^ was fitted using the lifelines Python library.^53^ Models included standardized baseline protein abundance, baseline age, sex, baseline CDR category, study site and PC1–PC5. Hazard ratios are reported per 1 s.d. higher baseline protein. Multiple testing was controlled using Benjamini–Hochberg FDR correction within each analysis scope across the 30 signature aptamers, with the CN+MCI scope reported as primary. As a sensitivity analysis, the CDR-worsening Cox model was additionally adjusted for *APOE ε*2 and *ε*4 allele dose, each coded as the number of copies (0, 1 or 2), in the genotyped subset.

### Stanford ADRC plasma biomarker association analysis

To test whether the 30-aptamer signature aligned with AD-related biomarker burden, we analyzed plasma protein and biomarker measurements from the Stanford Alzheimer’s Disease Research Center (Stanford ADRC), one of the contributing sites within GNPC. Stanford ADRC plasma samples were profiled on the SomaScan v4.1 platform and preprocessed using the same general pipeline applied to GNPC plasma, including log2 transformation, IQR-based outlier masking and k-nearest-neighbor imputation. Strict aptamer-level matching recovered 29 of the 30 discovery aptamers in Stanford ADRC; one aptamer was absent and was excluded.

We evaluated three core AD biomarkers, A*β*42/40,^13,14^ pTau-217^15,38^ and pTau-181,^16^ together with two additional AD-related injury and glial biomarkers, neurofilament light chain (NfL) and glial fibrillary acidic protein (GFAP). The A*β* 42/40 ratio was inverse-coded so that positive aligned effects represented the AD-pathology direction for all five biomarkers. For each of the 29 aptamers and five biomarkers, we fitted a linear mixed-effects model with the standardized biomarker as the outcome and standardized protein abundance as the predictor, adjusting for age, sex and the first five Stanford ADRC proteomic principal components, with a participant-level random intercept to accommodate repeated visits per participant, mirroring the mixed-effects framework used in the discovery analysis. Across the 145 protein–biomarker pairs, a median of 534 observations from 293–302 unique participants contributed to each model.

Multiple testing was controlled using a single Benjamini–Hochberg FDR family across all 145 protein–biomarker tests. Per-biomarker FDR values were also reported as sensitivity results in Supplementary Table 5. Direction concordance was assessed relative to the Q1 or Q3 assignment from the GNPC discovery model, accounting for the inverse coding of A*β*42/40.

### GNPC CSF concordance analysis

To test whether the directional AD-associated plasma signature was preserved in CSF, we performed a separate cross-tissue concordance analysis using GNPC CSF SomaScan v4.1 profiles. To mirror the plasma discovery contrast, the CSF analysis was restricted to CN and AD participants, excluding MCI participants. Because longitudinal CSF sampling was limited, the analysis used each participant’s first available CSF sample, yielding a cross-sectional CSF subset of 609 participants (323 controls and 286 AD).

CSF aptamer profiles were processed using a pipeline parallel to the plasma preprocessing workflow, including log2 transformation, IQR-based outlier masking, call-rate filtering, k-nearest-neighbor imputation and principal component analysis on the imputed CSF matrix. For each of 7,351 CSF aptamers, we fitted a cross-sectional linear model with CSF protein abundance as the outcome and AD status as the primary predictor, adjusting for sex, baseline age, study site and the first five CSF proteomic principal components. Controls were used as the reference group, so the AD coefficient represented the cross-sectional AD-versus-control difference in CSF abundance.

Benjamini–Hochberg FDR correction was applied to the AD-term *p* values across all 7,351 CSF aptamers. This analysis was used as a cross-tissue concordance domain rather than as a new discovery step: plasma signature aptamers were evaluated for whether they retained a direction-consistent AD association in CSF at FDR < 0.05.

### UK Biobank Olink replication of incident AD and dementia

UKB replication used the Olink Explore panel from the UK Biobank Pharma Proteomics Project (UKB-PPP), which profiled 2,923 plasma proteins in 54,219 participants.^21^ We used the preprocessed, imputed UKB Olink protein matrix generated under the quality-control framework described in,^54^ yielding 44,526 participants with protein measurements and complete covariates for survival modeling. Participants with prevalent disease at baseline were then excluded separately for each outcome, leaving 38,243 participants for incident AD (622 events) and 38,213 for incident all-cause dementia (1,171 events).

Of the 30 signature aptamers, 18 unique genes had a corresponding Olink analyte under gene-symbol matching and were tested. Participants were followed from baseline age to outcome onset age or last known age. For each protein and outcome, a Cox proportional hazards model was fitted with lifelines, including standardized protein abundance, baseline age, sex and the first five Olink proteomic principal components.

Multiple testing was controlled by a single Benjamini–Hochberg FDR family across 18 proteins × 2 outcomes (36 tests). Associations with FDR < 0.05 and direction consistent with the GNPC discovery quadrant were considered supportive external replication.

### Plasma pQTL Mendelian randomization

To provide orthogonal genetic support for the longitudinal plasma signature, we performed two-sample Mendelian randomization (MR) using plasma protein quantitative trait loci (pQTLs) as exposure instruments and AD GWAS summary statistics as the outcome.^55–57^ Plasma pQTL instruments were obtained from two large proteogenomic resources: the deCODE SomaScan plasma study^20^ and the UKB-PPP Olink platform.^21^

For each protein, candidate cis-pQTL instruments were selected from variants associated with plasma protein abundance, present in the AD GWAS outcome dataset, and meeting minor allele frequency ≥ 0.01. To reduce redundancy from correlated regional signals, instruments were restricted to at most one variant per 1 Mb region per protein. Highly pleiotropic variants were excluded, with pleiotropy defined as association with more than three proteins across the instrument-selection framework.

Exposure and outcome datasets were harmonized by effect allele before MR analysis. For deCODE SomaScan, instruments were matched to GNPC signature proteins by exact SomaScan aptamer identifier, with no gene-level fallback because different aptamers targeting the same gene may capture distinct epitopes or isoforms. For UKB-PPP Olink, gene-level matching was used because the Olink resource reports one measurement per gene. Primary MR estimates were defined as inverse-variance weighted estimates when at least two instruments were available and Wald-ratio estimates when only one instrument was available. MR-Egger, weighted-median and Cauchy estimates were retained in the raw output but were not used for primary evidence classification.

Benjamini–Hochberg FDR correction was applied separately within each plasma pQTL source among signature proteins with matched instruments. Unmatched signature proteins were coded as unavailable and were not included in the source-specific FDR family.

For any signature protein reaching FDR < 0.05 in plasma pQTL MR, we tested whether the cis-pQTL and AD GWAS associations shared a causal variant, since an MR estimate based on few cis instruments can reflect linkage disequilibrium rather than a shared causal signal. Colocalization was assessed with the approximate Bayes factor method in coloc,^58^ using all variants present in both the cis-pQTL and AD GWAS datasets across the ±1 Mb cis window, with default priors (*p*_1_ = *p*_2_ = 10^-4^, *p*_12_ = 10^-5^). Support for a shared causal variant was read from the posterior probability of the colocalization hypothesis (PP.H4).

### Rare-variant loss-of-function burden

As a second genetic line of evidence, we performed a gene-based rare-variant burden meta-analysis of Alzheimer’s disease and related dementias (ADRD) and looked up all 30 signature genes. Within each dataset, rare variants (minor allele frequency < 1%) were collapsed per gene and tested for association with ADRD case status using REGENIE^59^ gene-based logistic burden models with Firth correction, adjusting for genetic principal components, sex, age and APOE ε2 and ε4 dosage. Burden tests from whole-genome sequencing in UKB,^22^ the All of Us (AoU) Research Program^23^ and the Alzheimer’s Disease Sequencing Project (ADSP)^24^ were combined with previously published whole-exome-sequencing ADRD case–control summary statistics (Holstege et al.^25^) by inverse-variance-weighted fixed-effects meta-analysis, restricted within each dataset to genes with a cumulative minor allele count ≥ 10; depending on this threshold, 1 to 4 datasets contributed to each gene. The European-ancestry meta-analysis (up to 522,783 individuals: UKB 343,362, AoU 134,925, ADSP 23,151 and Holstege 21,345) was taken as primary, to match the European pQTL and GWAS resources used for MR, with the multi-ancestry meta-analysis reported alongside it. The clinically ascertained proband ADRD meta-analysis was used as the primary phenotype; by-proxy case definitions, which dilute case status, were not used. Variants were annotated with the Ensembl Variant Effect Predictor (VEP v114)^60^ against the GRCh38 reference. Loss-of-function status was assigned by LOFTEE^61^ and allele frequencies were taken from gnomAD v4.1.^62^ Multiallelic sites were decomposed into biallelic variants, and analysis was restricted to protein-coding genes. High-confidence loss-of-function (LoF) variants, defined as those with a HIGH-impact consequence carrying a LOFTEE high-confidence flag, formed the variant set.

For each gene we recorded the burden odds ratio and p value. Because rare LoF variants reduce gene product, a burden estimate was defined as direction-concordant when a risk-increasing odds ratio (OR > 1) accompanied a protein that declines in AD (Q3) and a risk-decreasing odds ratio (OR < 1) accompanied a protein that rises in AD (Q1). Benjamini–Hochberg FDR was computed within the 30-gene family per ancestry. Genes with a direction-concordant burden at p < 0.05 were reported as nominal genetic support, and signals attributable to a single contributing cohort were flagged. A gene was counted as supported in the genetic domain if it reached direction-concordant significance in either plasma pQTL MR or rare-variant burden.

## Data availability

GNPC plasma and CSF SomaScan v4.1 data are available to qualified researchers through the Alzheimer’s Disease Data Initiative AD Workbench under a Data Use Agreement. UK Biobank Olink proteomic data are available via the UK Biobank application process. Stanford ADRC data are available from the Stanford ADRC investigators on reasonable request, subject to institutional data-sharing policies. deCODE and UK Biobank Pharma Proteomics Project plasma pQTL summary statistics, and the Alzheimer’s disease GWAS summary statistics used as the Mendelian randomization outcome, are publicly available from the respective consortia. The rare-variant burden meta-analysis used exome and genome sequencing from UK Biobank, the All of Us Research Program and the Alzheimer’s Disease Sequencing Project (ADSP), each accessible to approved researchers through the respective program data-access procedures. Previously published ADRD case–control burden summary statistics (Holstege et al.) are available at https://doi.org/10.5281/zenodo.6818051.

## Code availability

Analysis code is available at https://github.com/Junkkkk/gnpc-longitudinal-ad-proteomics.

## Acknowledgments

We thank the Global Neurodegeneration Proteomics Consortium (GNPC) for the biospecimens, clinical data and proteomic data that made this work possible. The harmonized GNPC dataset was accessed and analyzed through the Alzheimer’s Disease Data Initiative AD Workbench, and data discovery and analysis services were provided in-kind by the AD Data Initiative. The majority of GNPC biosample proteomic analyses were supported by Gates Ventures and Johnson & Johnson, as described in the GNPC resource publication.^11^ This research was conducted using the UK Biobank Resource under Application Number 45420. We thank all the participants and researchers of UK Biobank for making these data available. This work was supported in part by the National Institutes of Health (P30 AG066515).

## Author contributions

J.P. designed the study, performed the discovery and downstream evidence analyses, generated the figures and wrote the manuscript. Y.L.G. designed and performed the Mendelian randomization and rare-variant burden analyses. M.D.G. supervised the study. All authors provided critical edits and reviews of the manuscript.

## Competing interests

The authors declare no competing interests.

## Supplementary Information Supplementary Figures

### Supplementary Figures

**Supplementary Figure 1.**
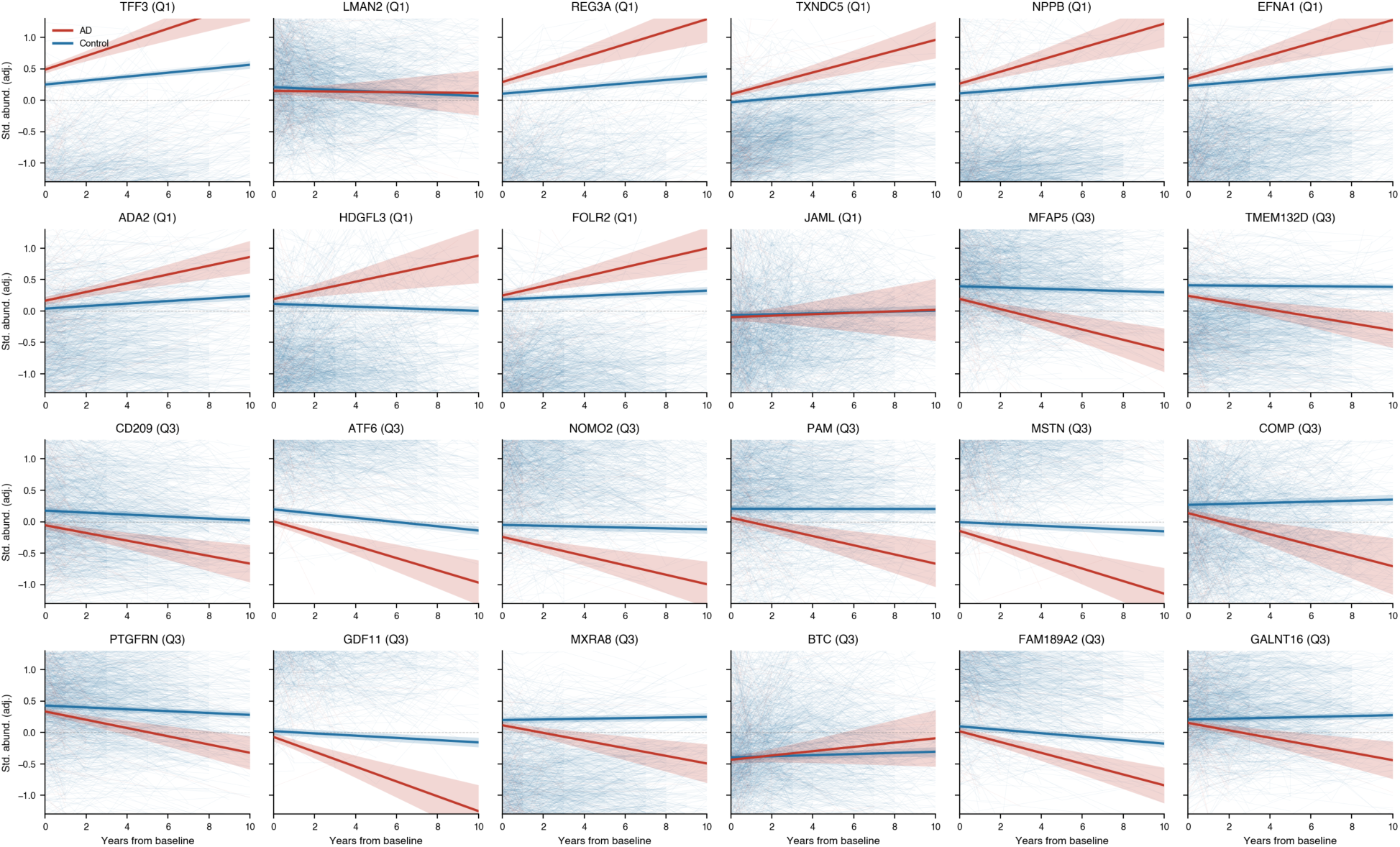
Covariate-adjusted longitudinal trajectories for the 24 signature proteins not shown in. Figure 2d **(10 Q1, 14 Q3).** For each protein, log2 abundance was standardized (z-scored) and adjusted for sex, baseline age, study site and the first five plasma proteomic principal components. Bold lines show model-predicted mean trajectories for AD (red) and control (blue) at mean covariate values, with shaded 95% confidence bands; faint lines show individual covariate-adjusted trajectories for participants with two or more plasma visits. Q1 proteins rose and Q3 proteins declined in AD relative to controls over follow-up, while control trajectories stayed near-flat. Trajectories are shown to 10 years, beyond which fewer than 6% of samples contributed.

## Supplementary Tables

**Supplementary Table 1.** Full GNPC plasma longitudinal mixed-effects results. For each of the 7,362 SomaScan v4.1 aptamers, baseline AD main-effect and AD-by-time interaction estimates (*β*, standard error and *P*) are reported in long format (one row per aptamer and term), with per-term Benjamini–Hochberg FDR computed separately for each term across all aptamers.

**Supplementary Table 2.** Discovery sensitivity analyses. For each of the 7,362 SomaScan aptamers, baseline (AD main effect) and AD-by-time interaction estimates with nominal *p* values and per-term FDR are reported for the primary site-adjusted discovery model and for four alternative models; aptamers belonging to the 30-protein signature are flagged. Models: no site adjustment (*n* = 8,730; signature 87, of which 29 of the primary 30 were retained); measured-CDR baseline diagnosis (*n* = 6,290; signature 39); sites contributing both control and AD participants (*n* = 7,529; signature 27); and *APOE ε*2/*ε*4 allele-dose adjustment as a confounding control (*n* = 6,313; signature 39). Across all four models the 30 signature proteins remained concordant in direction on both terms (30/30), and effect-size estimates were highly correlated with the primary model (Pearson *r* = 0.985, 0.990, 0.999 and 0.997 for the baseline effect and 0.999, 0.898, 0.997 and 0.987 for the interaction, respectively); the smaller number re-clearing dual-term FDR in the reduced-sample models reflects statistical power rather than loss of signal.

**Supplementary Table 3.** Cross-domain evidence matrix for the longitudinal AD plasma signature. Evidence-domain results are summarized for all 30 signature proteins across clinical progression, Stanford ADRC plasma biomarkers, GNPC CSF, UK Biobank prospective replication and genetic support. Per-domain effect sizes, *P* values, FDR values and the direction-consistency or support flags used to define the multi-domain subset are included.

**Supplementary Table 4.** GNPC CDR-worsening Cox models for the 30-protein longitudinal AD plasma signature, across three diagnostic scopes (CN+MCI primary, CN-only, and CN+MCI+AD). For each protein and scope, the hazard ratio per 1 s.d. higher baseline abundance, 95% confidence interval, *p* value and per-scope FDR are reported. As a sensitivity analysis, hazard ratios from the genotyped subset without and with adjustment for *APOE ε*2/*ε*4 allele dose, and their ratio, are also given.

**Supplementary Table 5.** Stanford ADRC plasma biomarker associations. Linear mixed-effects association results are shown for the 29 signature aptamers assayed in Stanford ADRC across five AD-related plasma biomarkers (A*β*42/40, pTau-181, pTau-217, GFAP and NfL). Effect sizes, *P* values, FDR values and direction-consistency flags are reported across the combined family of 145 protein–biomarker tests.

**Supplementary Table 6.** UKB Olink prospective replication. Cox proportional hazards results are shown for the 18 signature proteins assayed on UKB Olink across incident AD and all-cause dementia. Hazard ratios, 95% confidence intervals, *P* values and FDR values are reported with Benjamini–Hochberg correction across the 36 protein–outcome tests.

**Supplementary Table 7.** GNPC CSF concordance analysis. Cross-sectional CSF association results (AD versus cognitively normal controls) for the 30 plasma signature proteins. Effect sizes, *P* values, per-protein Benjamini–Hochberg FDR (computed across all GNPC CSF aptamers) and plasma-signature direction-concordance flags are reported.

**Supplementary Table 8.** Genetic support for the 30-protein longitudinal AD plasma signature: plasma pQTL Mendelian randomization (deCODE and UKB-PPP) with *ANTXR1* colocalization. Inverse-variance weighted MR estimates with per-source Benjamini–Hochberg are reported.

**Supplementary Table 9.** Rare-variant loss-of-function burden for the 30 longitudinal AD plasma signature genes. Gene-based high-confidence loss-of-function burden associations from an ADRD proband meta-analysis are reported for the European-ancestry analysis, with odds ratios, p-values, Benjamini–Hochberg FDR computed within the 30-gene family, per-cohort effect direction, direction concordance, and nominal and FDR significance flags.

**Supplementary Table 10.** Repeated-sample structure of the GNPC plasma discovery cohort by contributing site and baseline diagnosis. For each site, the number of AD and control participants and their corresponding longitudinal sample counts are tabulated, together with site totals. This table documents the multi-visit structure underlying the longitudinal mixed-effects discovery analysis.

## Notes

### Competing Interest Statement

The authors have declared no competing interest.

### Author Declarations

Ethics committees or Institutional Review Boards of the contributing institutions in the Global Neurodegeneration Proteomics Consortium gave ethical approval for the original studies, and written informed consent was obtained from participants or their legal representatives at the site of origin. This study used de-identified, harmonized data released through the Global Neurodegeneration Proteomics Consortium Health Data Stewardship framework and accessed through the Alzheimer's Disease Data Initiative AD Workbench under the Global Neurodegeneration Proteomics Consortium Data Use Agreement and publication policies. UK Biobank has ethical approval from the North West Multi-centre Research Ethics Committee, and all participants provided written informed consent. UK Biobank data were accessed under an approved UK Biobank application.

